# Clinical Characteristics and Treatment Experience of Individuals with SCN8A Developmental and Epileptic Encephalopathy (SCN8A-DEE): Findings from an Online Caregiver Survey

**DOI:** 10.1101/2021.11.29.21267027

**Authors:** Alison Cutts, Hillary Savoie, Michael F. Hammer, John Schreiber, Celene Grayson, Constanza Luzon, Noam Butterfield, Simon N. Pimstone, Ernesto Aycardi, Cynthia Harden, Chuck Yonan, Eric Jen, Trung Nguyen, Tara Carmack, Dietrich Haubenberger

**Affiliations:** Xenon Pharmaceuticals Inc., Burnaby, BC, Canada; The Cute Syndrome Foundation, Troy, NY, USA; Shay Emma Hammer Research Foundation and University of Arizona, Tucson, AZ, USA; Children’s National Hospital, Washington DC, USA; Neurocrine Biosciences, Inc., San Diego, CA, USA

**Keywords:** SCN8A-DEE, SCN8A-related epilepsy, caregiver survey, seizures, phenotype, treatment

## Abstract

**Purpose:** SCN8A developmental epileptic encephalopathy (SCN8A-DEE) is a rare and severe genetic epilepsy syndrome characterized by early-onset developmental delay, cognitive impairment, and intractable seizures. Variants in the SCN8A gene are associated with a broad phenotypic spectrum and variable disease severity. A caregiver survey, solicited by the advocacy group The Cute Syndrome Foundation (TCSF), was conducted to gather information on the demographics/disease presentation, seizure history, and treatment of patients with SCN8A-related epilepsies.

**Methods:** A 36-question online survey was developed to obtain de-identified data from caregivers of children with SCN8A-related epilepsy. The survey included questions on genetic diagnosis, disease manifestations/comorbidities, seizure severity/type, current/prior use of antiseizure medicines (ASMs), and best/worst treatments per caregiver perception.

**Results:** In total, 116 survey responses (87 USA, 12 Canada, 12 UK, 5 Australia) were included in the quantitative analysis. Generalized tonic/clonic was the most common seizure type at onset and time of survey; absence and partial/focal seizures were also common. Most patients (77%) were currently taking ≥2 ASMs; 50% had previously tried and stopped ≥4 ASMs. Sodium channel blockers (oxcarbazepine, phenytoin, lamotrigine) provided the best subjective seizure control and quality of life.

**Conclusion:** The SCN8A-DEE patient population is heterogeneous and difficult to treat, with high seizure burden and multiple comorbidities. The high proportion of patients who previously tried and stopped ASMs indicates a large unmet treatment need. Further collaboration between families, caregivers, patient advocates, clinicians, researchers, and industry can increase awareness and understanding of SCN8A-related epilepsies, improve clinical trial design, and potentially improve patient outcomes.

**HIGHLIGHTS:**

- This is the first survey-based study of caregiver experiences in SCN8A-DEE
- Caregivers report a broad range of seizure types and genetic variants in patients
- Patients generally suffer from high seizure burden and multiple comorbidities
- Results suggest new treatments and standardized treatment protocols are needed
- Patient-centered research may improve awareness of SCN8A-DEE and patient outcomes

## INTRODUCTION

SCN8A developmental and epileptic encephalopathy (SCN8A-DEE) is a rare and serious genetic epilepsy syndrome characterized by early-onset developmental delay and regression, cognitive impairment, and multiple intractable seizure types.^1-3^ The SCN8A gene is highly expressed in the central nervous system and encodes the Na_v_1.6 voltage-gated sodium ion channel, which has a major role in regulating the excitatory neuronal networks in the brain.^1-3^

The first human epileptogenetic SCN8A mutation to be identified was a missense gain-of-function mutation that altered the biophysical properties of the Na_v_1.6 sodium channel, as reported by Veeramah et al. in 2012.^4-6^ Subsequent studies have confirmed the critical role of SCN8A in the initiation and propagation of action potentials and neuronal activity in the brain, and almost 400 individuals with SCN8A-related epilepsy have been described in the literature or documented in registries.^1-3,5,7-10^ The majority of documented SCN8A mutations are de novo gain-of-function mutations that result in premature channel opening, impaired inactivation, or elevated persistent current.^1-3,5,7^ Loss-of-function variants associated with a reduction of peak current have also been reported and are typically characterized by less severe phenotypes, including less severe developmental delay, cognitive impairment, movement disorder (e.g., myoclonus), and mild or no seizures.^1,11,12^

Although rapid progress continues to be made in the functional and phenotypic classification of SCN8A-related epilepsy, an understanding of its clinical presentation, progression, prognosis, and impact is just beginning to emerge.^1-3^ Pathogenic SCN8A mutations have been associated with a broad phenotypic spectrum and varying degrees of disease severity, ranging from movement disorders or intellectual disability (with or without seizures) to severe DEE.^1-3,13-16^ Individuals with less severe or “intermediate” DEE may have extended seizure-free periods, mild-to-moderate developmental delay, and mild or absent neurological deficits, while those with the most severe DEE tend to have multiple seizure types which do not or only insufficiently respond to treatment, severe and progressive physical and cognitive impairment, and increased rates of early mortality.^1-3,13-15,17^

The most common seizure type in individuals with SCN8A-related epilepsies is generalized tonic/clonic, although partial/focal, absence, myoclonic, and atonic seizures have also been reported.^18^ Seizures typically manifest within the first 18 months after birth, with an average age at seizure onset of 4 months.^1-3^ Electroencephalographic (EEG) findings are variable and can include a slow background and generalized, frontal, temporo-occipital, and multifocal epileptiform discharges.^2,18^ Additional clinical features of SCN8A-related epilepsies are heterogeneous and may include pyramidal and extrapyramidal movement disorders (e.g., myoclonus, dystonia, dyskinesia, and choreoathetosis), hypotonia, autism, progressive cortical visual impairment, and gastrointestinal disorders.^1,2,13^ About half of children with SCN8A-DEE do not develop the ability to walk or sit properly, and patients have an increased risk of death from multiple causes, including sudden unexpected death in epilepsy (SUDEP).^3,13,18^ Published data on the treatment experiences of individuals with SCN8A-DEE and SCN8A-related epilepsies are limited, but commonly indicate that in most cases of severe SCN8A-DEE, seizures are extremely drug-resistant.^3^ Some studies have reported that patients achieved a certain level of seizure reduction or seizure-free periods with sodium channel blockers such as phenytoin, carbamazepine, oxcarbazepine, and valproate.^13,18^ However, these non-selective sodium channel blockers must often be administered at supratherapeutic doses, which can lead to poor tolerability.^13,18^ Other treatments such as lamotrigine and topiramate have shown poor or inconsistent effect,^13,18^ while treatment with levetiracetam has been reported to sometimes worsen seizures in patients with SCN8A-related epilepsy.^19^

Previous studies characterizing the heterogeneity of clinical presentation, progression, and prognosis of SCN8A-DEE and SCN8A-related epilepsies have been based on clinic samples with limited sample sizes. This international online caregiver survey was conducted to gather information on the demographics and disease presentation, seizure history, and treatment of individuals with this rare condition. These survey results provide insight into the patient’s treatment experience and add to the existing literature from a patient perspective.^10^

## METHODS

### Survey Procedure

This study was a cross-sectional study design. A 36-question online survey was developed by Xenon Pharmaceuticals, Inc. in collaboration with The Cute Syndrome Foundation (TCSF), an advocacy group that raises awareness of SCN8A-related epilepsies, works with those focused on treating this disorder, and provides support for families of children with this disorder. The survey was organized into sections on demographics and diagnosis, seizure history, seizure treatment, and future clinical trials, and included questions about genetic diagnosis, disease manifestations and comorbidities, seizure severity and type, current and prior use of antiseizure medicines (ASMs), as well as best and worst treatments per caregiver perception. Survey questions were either multiple choice (select one answer or select all that apply) or open-ended (allowing for free-text responses). The survey design and materials were approved by an Institutional Review Board and caregivers provided informed consent before completing the survey. The complete survey is included in the Supplementary Materials.

### Participants

Caregivers of children with SCN8A-related epilepsy were recruited to participate in the online survey over a 3-week period in late 2019 via targeted geography (United States, Canada, United Kingdom, and Australia only), targeted email outreach, a social media campaign, and an educational webinar. The survey was conducted by invitation only. Participation was voluntary, but respondents were offered a small financial compensation for their time in completing the survey. Duplicate caregiver responses, responses from non-English-speaking countries, or responses from outside the jurisdiction of the Institutional Review Board were excluded from the analysis. To avoid analyzing ambiguous data (e.g., responses for current seizure frequency or current ASMs), caregiver responses were not included in the quantitative analysis if the patient with SCN8A-related epilepsy was deceased at the time of the survey.

### Analysis

Survey responses were collected and collated over a 3-week period in late 2019. Survey results were analyzed descriptively, and free-text responses were manually reviewed and grouped according to themes. Odds ratios and association tests between quality of life and seizure control for antiepileptic treatments were determined by chi-square analysis or Fisher’s exact test.

## RESULTS

### Participants

A total of 161 caregivers of patients with SCN8A-related epilepsies were invited to take part in the survey. Of 125 survey responses received, 3 were excluded (living in non-English speaking country, n=2; duplicate responses, n=1). Additionally, 6 responses were received from caregivers of deceased patients and these responses were not included in the quantitative analysis. The remaining 116 responses were included in the analysis. The majority of caregiver respondents (75%) were from the United States; the others were from the United Kingdom, Canada, and Australia (**Table 1**). The majority of patients (n=76) were aged 2 to <12 years at the time of survey (current). There were only 7 patients aged ≥18 years at survey collection. The mean/median age at seizure onset was 0.75/0.33 years, with a range of 0 to 13 years. A total of 74 SCN8A variants were reported in the survey, the most common of which were R850Q (n=6), G1475R (n=6), and R1872W (n=4). Other variants found in ≥2 individuals were A1323T, R1617Q, N1877S (n=3 for each), and R223S, Q417P, L977P, G1451S, R1872L (n=2 for each). The remaining 63 variants were each reported in single patients only. Most of these variants have been previously described in patients with SCN8A-related epilepsy: R850Q,^13,19^ G1475R,^13,14,19^ R1872W,^13,14,19^ A1323T,^19,20^ R1617Q,^13,14,19^ N1877S,^13,19^ G1451S,^19^ and R1872L.^13,19^ Several of these variants have also been characterized as gain-of-function in functional studies: G1475R,^21,22^ R1872W,^7,21-23^ R1617Q,^7^ and R1872L.^7,22,23^

**Table 1.** Patient Characteristics by Current Age.

|  | **0 to <2 Years**<br>**(n=13)** | **2 to <12**<br>**Years**<br>**(n=76)** | **12 to <18**<br>**Years**<br>**(n=20)** | **18 Years or**<br>**Older**<br>**(n=7)** | **Overall**<br>**Study**<br>**Population**<br>**(N=116)** |
| --- | --- | --- | --- | --- | --- |
| **Country of residence, n (%)** |  |  |  |  |  |
| Australia | 0 (0) | 2 (3) | 3 (15) | 0 (0) | 5 (4) |
| Canada | 0 (0) | 11 (14) | 1 (5) | 0 (0) | 12 (10) |
| United Kingdom | 1 (8) | 7 (9) | 3 (15) | 1 (14) | 12 (10) |
| United States | 12 (92) | 56 (74) | 13 (65) | 6 (86) | 87 (75) |
| **Age at seizure onset** |  |  |  |  |  |
| Mean (SD), years | 0.18 (0.15) | 0.54 (1.10) | 1.23 (2.73) | 2.75 (5.07) | 0.75 (1.91) |
| Median (range), years | 0.17<br>(0.01-0.58) | 0.33<br>(0-8) | 0.38<br>(0.08-12) | 0.58<br>(0-13) | 0.33<br>(0-13) |
| **Seizure frequency at onset, n (%)<sup>a</sup>** |  |  |  |  |  |
| >10 per day | 1 (8) | 9 (14) | 5 (25) | 0 (0) | 15 (14) |
| 2-10 per day | 6 (46) | 19 (29) | 6 (30) | 1 (20) | 32 (31) |
| 1 per day | 1 (8) | 10 (15) | 3 (15) | 0 (0) | 14 (13) |
| 1 per week | 4 (31) | 18 (27) | 3 (15) | 2 (40) | 27 (26) |
| 1 per month | 1 (8) | 5 (8) | 1 (5) | 1 (20) | 8 (8) |
| <1 per month | 0 (0) | 5 (8) | 2 (10) | 1 (20) | 8 (8) |
| **Current seizure frequency, n (%)<sup>b</sup>** |  |  |  |  |  |
| >10 per day | 3 (23) | 6 (9) | 2 (10) | 0 (0) | 11 (10) |
| 2-10 per day | 2 (15) | 5 (7) | 4 (20) | 2 (33) | 13 (12) |
| 1 per day | 1 (8) | 10 (14) | 0 (0) | 1 (17) | 12 (11) |
| 1 per week | 1 (8) | 8 (12) | 2 (10) | 2 (33) | 13 (12) |
| 1 per month | 1 (8) | 7 (10) | 2 (10) | 1 (17) | 11 (10) |
| <1 per month | 2 (15) | 12 (17) | 6 (30) | 0 (0) | 20 (19) |
| Seizure-free | 2 (15) | 19 (28) | 4 (20) | 0 (0) | 25 (23) |
| Unknown but not seizure-free | 1 (8) | 2 (3) | 0 (0) | 0 (0) | 3 (3) |
| **Longest seizure-free period in past 6 months, n (%)<sup>b</sup>** |  |  |  |  |  |
| 0 days | 3 (23) | 6 (9) | 2 (10) | 1 (17) | 12 (11) |
| 1 day to 2 weeks | 3 (23) | 17 (25) | 4 (20) | 4 (67) | 28 (26) |
| >2 weeks to 1 month | 3 (23) | 7 (10) | 5 (25) | 1 (17) | 16 (15) |
| >1 month to <3 months | 1 (8) | 11 (16) | 1 (5) | 0 (0) | 13 (12) |
| 3 to <6 months | 2 (15) | 9 (13) | 2 (10) | 0 (0) | 13 (12) |
| 6 months | 1 (8) | 19 (28) | 6 (30) | 0 (0) | 26 (24) |

<sup>a</sup> Based on available data: 0 to <2 years, n=13; 2 to <12 years, n=66; 12 to <18 years, n=20; 18 years or older, n=5; overall, n=104.
<sup>b</sup> Based on available data: 0 to <2 years, n=13; 2 to <12 years, n=69; 12 to <18 years, n=20; 18 years or older, n=6; overall, n=108.
SD, standard deviation.

### Disease Presentation and Severity

Initial seizure frequency was reported as ≥1 seizure per month in 92% of patients and ≥1 seizure per day in 59% of patients (**Table 1**). Overall seizure frequency decreased between onset and the time of the survey, with at least once-monthly seizures reported by 56% of respondents and once-daily seizures reported by 33% of respondents at the time of the survey. Additionally, 45% of patients initially experienced multiple seizures per day and 22% of patients were experiencing multiple seizures per day at the time of the survey. Within the last 6 months prior to survey date, caregivers of patients aged 2 to <18 years tended to report longer seizure-free periods (50.0% had periods of ≥1 month) compared to those of younger patients (30.8% had periods of ≥1 month). No patients 18 years or older had seizure-free periods of ≥1 month in the 6 months prior to survey date; however, there were only 7 patients aged ≥18 years at the time of survey. According to survey responses, a wide range of developmental and neurologic comorbidities were reported (**Table 2**). The most common of these were intellectual disability (74%), hypotonia (57%), and movement disorders (43%) such as ataxia, paroxysmal dyskinesia, dystonia, or choreoathetosis.

**Table 2.**
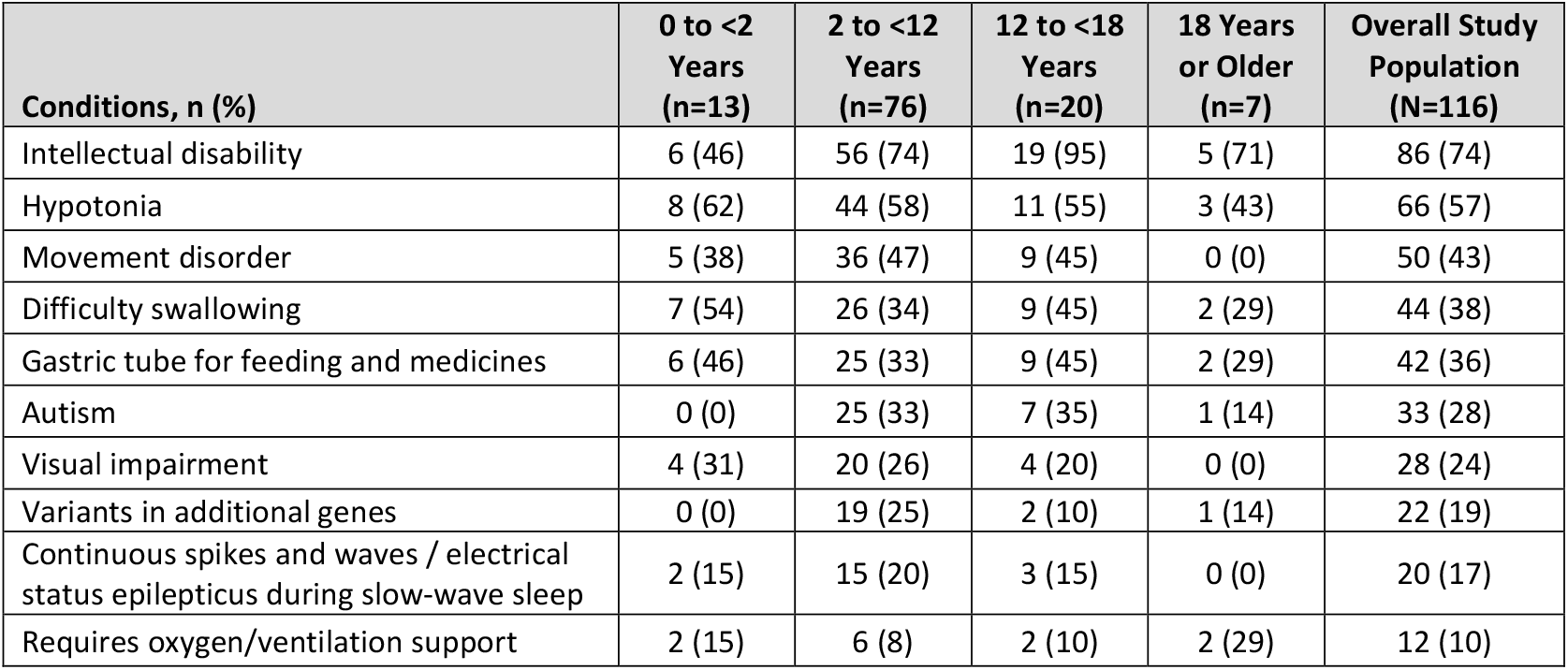
Comorbid Conditions by Current Age.

### Seizure Type and History

At seizure onset, generalized tonic/clonic was the most common seizure type reported by caregivers of patients <12 months of age (**Figure 1A**). Caregivers of patients aged ≥12 months at seizure onset reported a broader range of seizure types. At the time of survey (current), a broad range of seizure types was reported in all age groups (**Figure 1B**). Partial/focal was the most common current seizure type (73%) in patients <2 years of age. Generalized tonic/clonic was the most common seizure type in patients aged 2 to <12 years (62%) and 18 years and older (67%). Patients aged 12 to <18 years experienced a fairly even distribution of seizure types, with generalized tonic/clonic (44%), absence (44%), and partial/focal (38%) being the most common.

**Figure 1.**
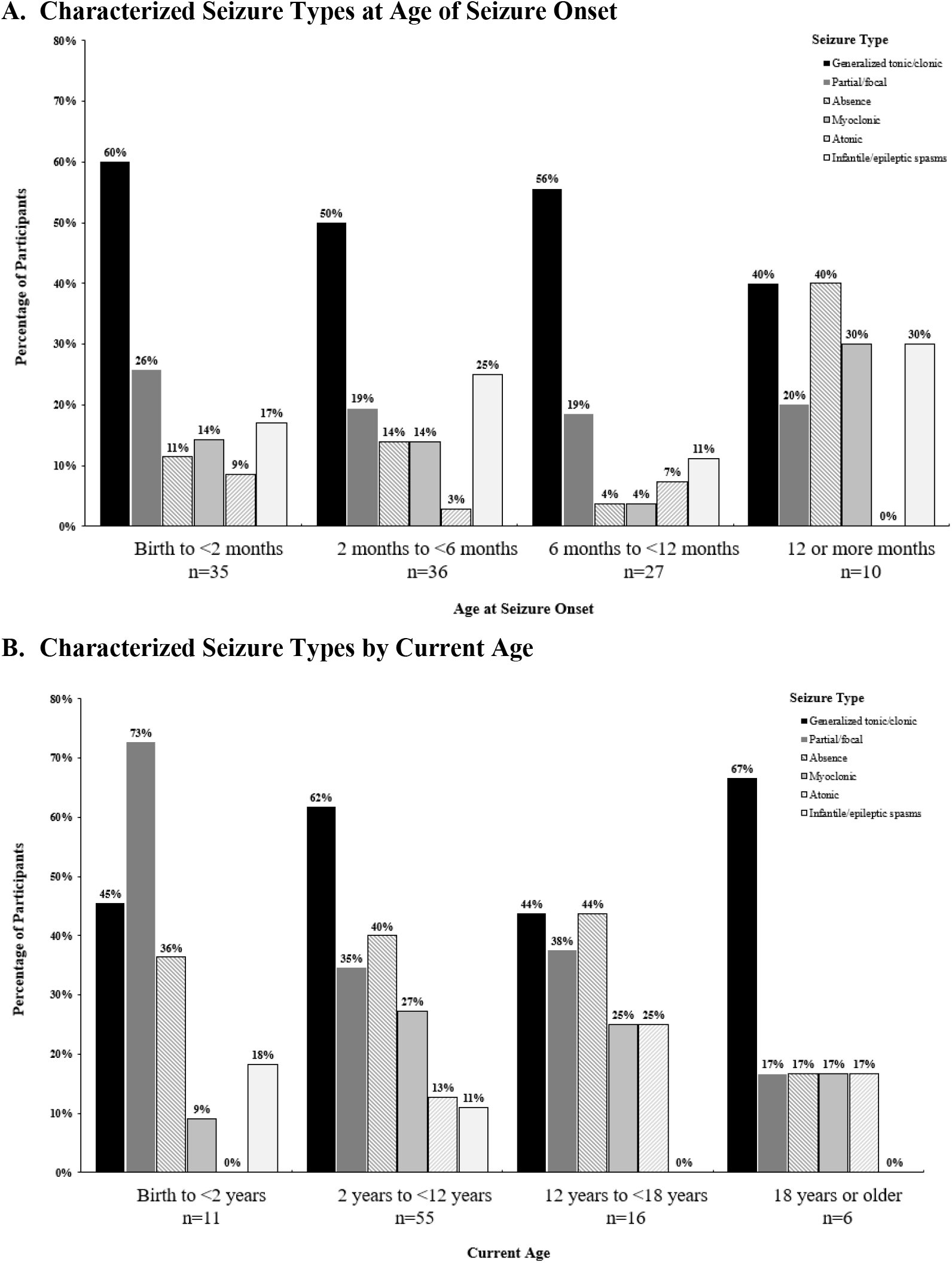
Seizure Types. Caregivers could report >1 seizure type per patient if needed.

Overall, 27% of patients were currently experiencing seizure clusters at least once per week, and 18% were experiencing daily seizure clusters (**Figure 2**). There were no consistent trends in terms of cluster frequency across the age groups, and most caregiver respondents did not provide information on the frequency of seizure clusters experienced in the past.

**Figure 2.**
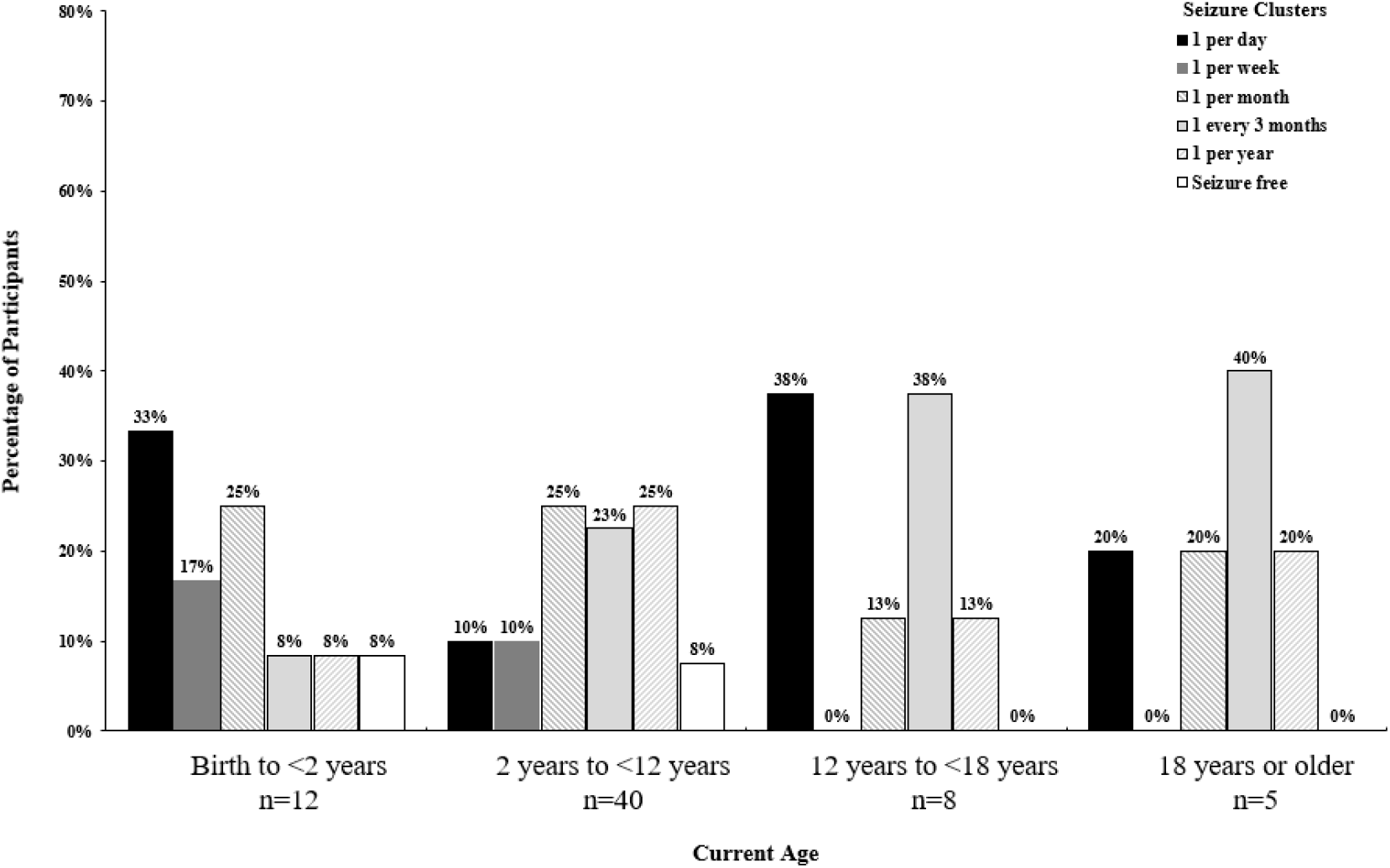
Current Seizure Cluster Frequency by Age.

### Seizure Treatment

The majority of patients (77%) were taking 2 or more ASMs, and almost a third (31%) of all patients were currently taking 4 or more ASMs. In total, 27% of patients were taking sodium channel blockers only, 9% of patients were taking other ASMs only, and 18% of patients were taking a combination of sodium channel blockers and other ASMs (**Table 3**). The majority of patients had tried and stopped 2 or more treatments and 50% had tried and stopped 4 or more treatments. Among those who reported what combination of ASMs they had taken or were currently taking, the most common combination therapy was oxcarbazepine and lacosamide (n=7). Lacosamide, followed by oxcarbazepine, was the most common ASM used in combination with any other ASM.

**Table 3.** Current ASM Treatments by Current Age^a^.

|  | **0 to <2**<br>**Years**<br>**(n=13)** | **2 to <12**<br>**Years**<br>**(n=76)** | **12 to <18**<br>**Years**<br>**(n=20)** | **18 Years or**<br>**Older**<br>**(n=7)** | **Overall Study**<br>**Population**<br>**(N=116)** |
| --- | --- | --- | --- | --- | --- |
| **Type of current ASM, n (%)<sup>b</sup>** |  |  |  |  |  |
| Sodium channel blocker (SCB) only | 1 (8) | 23 (30) | 5 (25) | 2 (29) | 31 (27) |
| Other ASM only (not SCB) | 1 (8) | 5 (7) | 4 (20) | 0 (0) | 10 (9) |
| Combination of SCB and other ASM | 6 (46) | 12 (16) | 2 (10) | 1 (14) | 21 (18) |
| Mixed/unknown MOA | 1 (8) | 5 (7) | 0 (0) | 0 (0) | 6 (5) |
| Not classified | 4 (31) | 31 (41) | 9 (45) | 4 (57) | 48 (41) |
| **Number of current ASMs, n (%)<sup>c</sup>** |  |  |  |  |  |
| 0 | 1 (8) | 1 (1) | 4 (20) | 0 (0) | 6 (6) |
| 1 | 1 (8) | 14 (20) | 4 (20) | 0 (0) | 19 (18) |
| 2 | 2 (15) | 14 (20) | 3 (15) | 0 (0) | 19 (18) |
| 3 | 5 (38) | 19 (28) | 4 (20) | 2 (33) | 30 (28) |
| ≥4 | 4 (31) | 21 (30) | 5 (25) | 4 (67) | 34 (31) |
| **Common current ASM combinations, n (%)<sup>d</sup>** |  |  |  |  |  |
| Oxcarbazepine/lacosamide | 2 (15) | 5 (7) | 0 (0) | 0 (0) | 7 (6) |
| Oxcarbazepine/lamotrigine | 0 (0) | 3 (4) | 0 (0) | 1 (14) | 4 (3) |
| Oxcarbazepine/phenytoin | 1 (8) | 0 (0) | 2 (10) | 0 (0) | 3 (3) |
| Clobazam/lamotrigine | 0 (0) | 3 (4) | 0 (0) | 0 (0) | 3 (3) |
| Lacosamide/phenytoin | 0 (0) | 1 (1) | 2 (10) | 0 (0) | 3 (3) |
| Lacosamide/clobazam/phenytoin | 0 (0) | 2 (3) | 0 (0) | 1 (14) | 3 (3) |
| **Number of ASMs tried and stopped, n (%)<sup>c</sup>** |  |  |  |  |  |
| 0 | 1 (8) | 10 (14) | 3 (15) | 1 (17) | 15 (14) |
| 1 | 2 (15) | 7 (10) | 4 (20) | 0 (0) | 13 (12) |
| 2 | 3 (23) | 7 (10) | 2 (10) | 1 (17) | 13 (12) |
| 3 | 1 (8) | 11 (16) | 0 (0) | 1 (17) | 13 (12) |
| ≥4 | 6 (46) | 34 (49) | 11 (45) | 3 (50) | 54 (50) |

<sup>a</sup> Based on available data: 0 to <2 years, n=13; 2 to <12 years, n=69; 12 to <18 years, n=20; 18 years or older, n=6; overall, n=108.
<sup>b</sup> Sodium channel blockers: carbamazepine, phenytoin, oxcarbazepine, lacosamide, and lamotrigine. Other ASMs: clonazepam, diazepam, ethosuximide, lorazepam, phenobarbital, primidone, gabapentin, tiagabine, vigabatrin, stiripentol, levetiracetam, and cannabidiol. Mixed/unknown MOA: rufinamide, valproate, felbamate, topiramate, and zonisamide. “Not classified” included patients receiving “miscellaneous” treatments not included in other categories (i.e., adrenocorticotrophic hormone, eslicarbazepine, brivaracetam, other cannabis products [e.g. Charlotte’s Web], dexamethasone, acetazolamide, clobazam, ketogenic diet/modified Atkins diet, sulthiame, ezogabine/retigabine, prednisone/prednisolone, pyridoxine/vitamin B6, and vagus nerve stimulator), patients with no drug use, and patients receiving combinations of drugs not otherwise noted (e.g., SCB + “miscellaneous”; SCB + “mixed/unknown MOA”, etc.).
<sup>c</sup> Including cannabis products, ketogenic diet, modified Atkins diet, and vagus nerve stimulator.
<sup>d</sup> Includes combinations reported in ≥3 patients.
ASM, antiseizure medicine; MOA, mechanism of action; SCB, sodium channel blocker.

At seizure onset, the most commonly prescribed ASMs were levetiracetam (n=51), phenobarbital (n=33), and topiramate (n=10). However, levetiracetam and topiramate were the 2 most commonly stopped ASMs (levetiracetam, n=54; topiramate, n=33), and were also reported to provide the worst seizure control (levetiracetam, n=35; topiramate, n=6) (**Figure 3A**). Free-text responses on reasons for worst or stopped ASMs are summarized in **Supplementary Table 1**. The most common reasons were lack of improvement, increase in seizure severity, and side effects. The most commonly reported side effects were gastrointestinal adverse effects, behavioral problems, and developmental delay.

**Figure 3.**
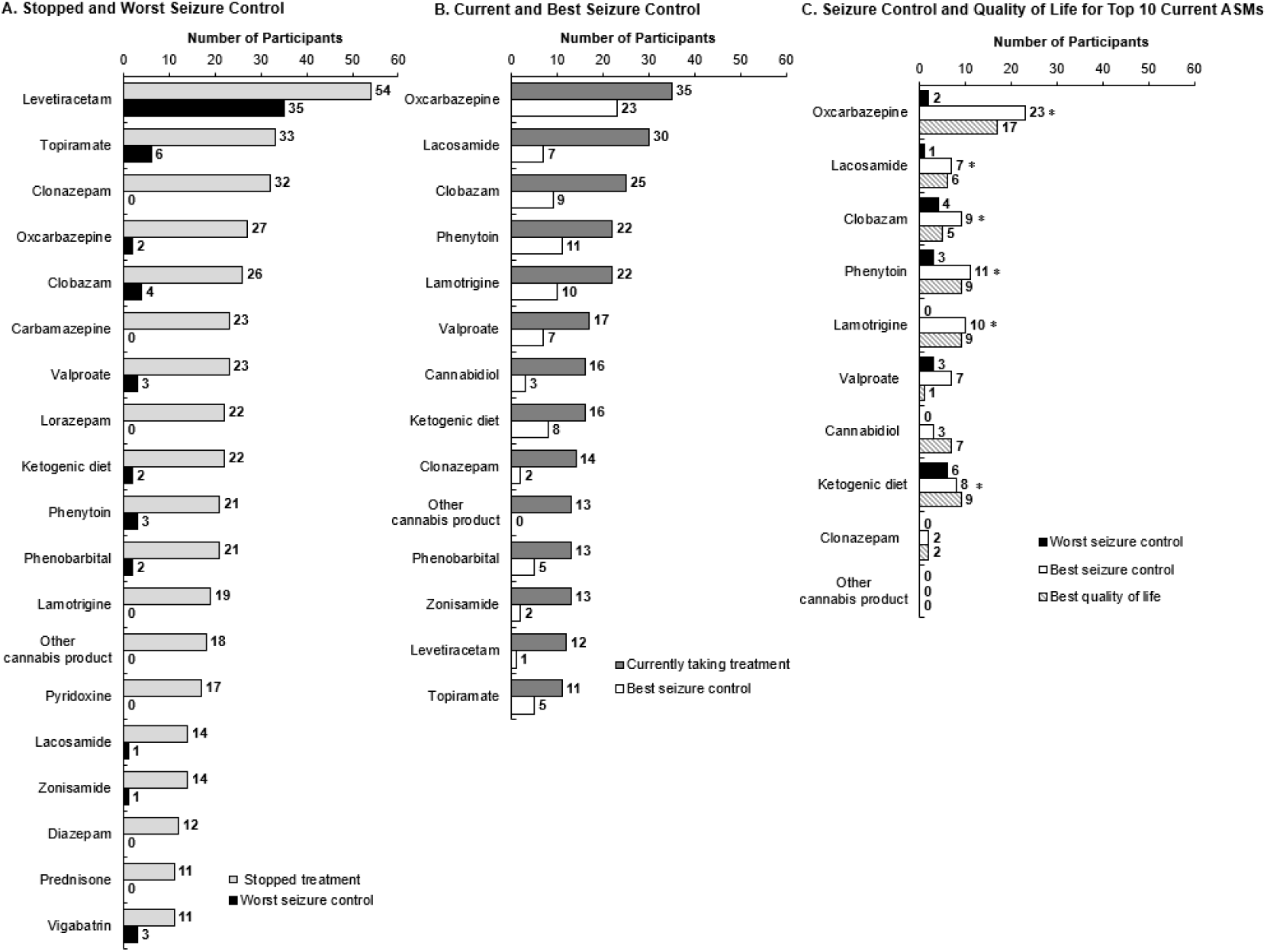
ASM Treatments and Caregiver Perceptions. Panels A and B include ASMs with >10 responses. * Compared to a response of “worst seizure control” (or no response), a response of “best seizure control” had significantly higher odds for a response of “best quality of life” (*P*<0.05). ASM, antiseizure medicine.

The most commonly used ASMs at time of the survey were oxcarbazepine (n=35), lacosamide (n=30), and clobazam (n=25) (**Figure 3B**). Oxcarbazepine, phenytoin, and lamotrigine were most commonly reported by caregiver respondents (per subjective perception) to provide the best seizure control (oxcarbazepine, n=23; phenytoin, n=11; lamotrigine, n=10) and best quality of life (oxcarbazepine, n=17; phenytoin, n=9; lamotrigine, n=9). Among the 10 most common current anti-seizure treatments, oxcarbazepine, lacosamide, clobazam, phenytoin, lamotrigine, and ketogenic diet had a significant correlation between responses for best seizure control and best quality of life (i.e., compared to a response of “worst seizure control” [or no response]. A response of “best seizure control” had significantly higher odds for a response of “best quality of life”, *P*<0.05) (**Figure 3C**).

## DISCUSSION

SCN8A-DEE and SCN8A-related epilepsies are extremely rare, having been described in almost 400 published cases to date.^8^ Existing characterizations of the heterogenous clinical presentation, progression, and prognosis of SCN8A-DEE and SCN8A-related epilepsies have been based on clinic samples with limited sample sizes. This is the first survey-based study of the experiences of caregivers of patients with SCN8A-related epilepsies. This study also provides valuable insight into the patient treatment experience, which has been inadequately described thus far. This study collected data from a substantial proportion of reported SCN8A-related epilepsy cases; over 77% (125/161) of contacted caregivers responded to the survey and 72% (116/161) were included in the analysis. The high response rate and multinational nature of this study give the findings of this study broad and global generalizability. This high response rate also highlights how thoughtful partnerships between advocacy organizations, industry, and academia can result in impressive response rates over short time periods. The high response rate may also be in part due to the efforts of patient advocates, who made a key contribution to the survey design by ensuring that the survey was patient-centered.

The results of this study suggest that the SCN8A-related epilepsy patient population is heterogenous, as caregivers reported a broad range of seizure types and genetic variants. As expected based on previous studies, caregivers reported a broad range of seizure types in patients both at onset and the time of survey, including generalized tonic/clonic, partial/focal, absence, myoclonic, and infantile/epileptic spasms.^7,13,14,19^ This study also found that generalized tonic/clonic seizures were especially common at seizure onset in patients under 12 months of age, which is consistent with previous reports.^4,7,15,24^ Caregivers also reported a broad range of genetic variants in this study, many of which were among the more than 250 SCN8A mutations that have been previously characterized.^8^ Repeat genetic mutations have been observed in about 25% of cases in previous studies^13^ compared to 50% in this study, perhaps in part to the increasing number of SCN8A mutations that have been characterized over time. Despite the heterogeneity of the patient population in this study, most patients suffered from a high seizure burden and multiple neurologic and motor comorbidities. The high proportion of caregiver respondents reporting daily seizures at onset (59%) and the time of survey (33%) matches previous reports,^19^ as does the high proportion of caregiver respondents reporting seizure clusters.^14^ Interestingly, caregivers in this study reported that seizure frequency tended to decrease from onset to the time of survey, seemingly contrary to the progressive worsening of epilepsy in most patients.^14^ This decrease in current seizure frequency relative to onset may be due to improving seizure control or the emergence of seizure-free periods (due to the refractory nature of SCN8A-related epilepsy) in some patients.^25^ The neurological comorbidities in this study also matched those reported previously, including the presence of intellectual disability, autistic features, motor disorders, gastrointestinal symptoms, and cortical blindness.^13^

Seizures in cases of severe SCN8A-DEE tend to be extremely drug resistant.^13,14^ This study found that a high proportion of patients previously tried and stopped various medications, with half of caregiver respondents reporting trying and stopping 4 or more ASMs. This high medication turnover suggests a general dissatisfaction with current therapies, which are likely inadequate for seizure control. Together, these findings suggest that there is an ongoing unmet need for effective and safe therapeutic options, as better seizure control could directly improve patients’ quality of life.

Caregivers of patients with SCN8A-related epilepsies reported using a wide range of ASMs at the time of the survey, with more than three in four patients taking >1 ASM and almost a third taking >4 ASMs. Such combination therapy is common in patients with SCN8A-related epilepsies.^3^ While no treatment guidelines or protocols for SCN8A-related epilepsies have been published, the efficacy of various treatments for SCN8A-related epilepsies has been characterized. For example, topiramate has been reported to have inconsistent effects,^13,14^ while levetiracetam can even exacerbate seizures.^13,14,19^ Caregiver respondents in this study also reported that topiramate and levetiracetam were the most frequently stopped ASMs and that levetiracetam resulted in the worst seizure control of all ASMs. Conversely, sodium channel blockers have previously shown inconsistent but beneficial effects in patients with SCN8A-related epilepsies,^13,14,19,26^ despite rare instances of worsening seizures.^13^ Specifically, oxcarbazepine, phenytoin, carbamazepine, and lamotrigine have been among the most effective sodium channel blockers,^13,14,19^ and improvement with zonisamide and lacosamide has also been reported.^13,26^ In this study, caregivers also reported sodium channel blockers (e.g., oxcarbazepine, lacosamide, phenytoin, and lamotrigine) to be among most effective ASMs. Like other ASMs, sodium channel blockers are often taken in combination with other therapies.^3^ Although sodium channel blockers were “pooled” for the purpose of this analysis, the ASMs reported to be most efficacious in this study (e.g., oxcarbazepine, lacosamide, phenytoin, and lamotrigine) act primarily through sodium channel blockade, as opposed to other sodium channel blockers with mixed mechanisms (e.g, valproic acid, topiramate, and zonisamide). Furthermore, there was a significant association between a response of “best seizure control” and “best quality of life” for these sodium channel blockers, suggesting sodium channel blockers improve patient and caregiver quality of life.

Understanding how genetic variants impact sodium channel function, seizure phenotype, and treatment response may allow clinicians to tailor treatments to individual patients’ needs. For example, the inconsistent response to sodium channel blockers in this and other studies may be due to loss-of-function SCN8A mutations in some patients. While typically associated with less severe phenotypes,^11,12^ loss-of-function SCN8A variations have also been described in individuals with SCN8A-related epilepsy.^24,27^ Genetic variants can also alter different electrophysiological properties of the Na_v_1.6 voltage-gated sodium ion channel, such as channel activation, inactivation, and persistent current,^21,22^ potentially resulting in different epilepsy phenotypes and treatment responses. Understanding these relationships may allow clinicians to increase the consistency of treatment response, thus improving patient outcomes. This study had several limitations. First, the retrospective nature of the study relies on the accuracy of caregiver recall, especially for the medications taken and seizure types at onset. For example, a recent study found that gain-of-function SCN8A mutations primarily caused focal epilepsy, suggesting that some of the generalized tonic/clonic seizures reported in this study could instead be classified as focal to bilateral tonic/clonic.^8^ Second, this study included a relatively small number of caregiver respondents and patients, especially patients aged 18 years or older. However, the 116 caregiver responses included in this study represent a large proportion of the total cases of SCN8A-related epilepsy described to date. Third, the high number of variants in other genes observed in this and previous studies^10^ introduces diagnostic uncertainty and suggests some patients may have had comorbidities that were unaccounted for in the survey.

In conclusion, the SCN8A-DEE patient population is heterogeneous and difficult to treat, suffering from a high seizure burden and multiple comorbidities. Despite rapid progress toward understanding the disease mechanism of SCN8A-related epilepsy and establishing better treatment protocols, there is a still large unmet treatment need and no standard treatment protocol in this patient population. The key contribution made by patient advocates to the survey design helped ensure that the survey was patient-centered, which can give caregivers and patients a greater voice in treatment development. To this end, the study was tailored to better understand seizure type, frequency, and burden with a goal of establishing clinical trial feasibility under various inclusion/exclusion scenarios. Unmet medical need was confirmed by caregiver responses, as 76.7% (89/116) of participants indicated a willingness to participate in future clinical trials. Cross-sectional studies like this one can complement retrospective chart review studies, prospective longitudinal registries, and natural history studies to assist in clinical trial design. Ultimately, collaboration between families, caregivers, patient advocates, clinicians, researchers, and industry can help increase awareness and understanding of this rare and serious disease, improve clinical trial design, and potentially improve overall patient outcomes.

## Data Availability

All data produced in the present work are contained in the manuscript

## Acknowledgements

Survey responses were collected by M3 Global Research and analyzed by Xenon Pharmaceuticals Inc. and Neurocrine Biosciences, Inc. We would also like to acknowledge all caregivers who participated in this study for their contributions.

## Supplementary Materials

### Survey

#### DEMOGRAPHICS and DIAGNOSIS

1. **Where do you live? To protect your privacy, please provide the approximate location only, do not provide your address or zip/postal code** Closest major city______________________*(free text box)* Country___________________*(free text box)*
2. **How old is your child?** *(select one)*
  1. Birth to less than 6 months
  2. 6 months to less than 1 year
  3. 1 year to less than IS months
  4. IS months to less than 2 years
  5. 2 years to less than 3 years
  6. 3 years to less than 4 years
  7. 4 years to less than 6 years
  8. 6 years to less than 12 years
  9. 12 years to less than 18 years
  10. 18 years and older
3. **Does your child have a genetic diagnosis of SCN8A-developmental and epileptic encephalopathy (SCN8A-DEE) or SCN8A-related epilepsy?**
  1. Yes, the doctor is sure they have SCN8A-DEE or SCN8A-related epilepsy *(answer question4)*
  2. No *(skip to question 6)*
  3. Unsure/Maybe *(skip to question 6)*
4. **Do you know what variant or mutation in SCN8Ayour child has?** *(select one)*
  1. Yes, I have a copy of my genetic testing report and the variant is (please provide)_______________*(free text box)*
  2. Yes, I don’t have a copy of my genetic testing report, but the doctor/genetic counsellor told me and the variant is (please provide)__________________*(free text box)*
  3. No, I don’t know the variant or mutation
5. **At approximately what age did your child receive their genetic diagnosis of SCN8A-DEE or SCN8A-related epilepsy? Please provide your best estimate in days, weeks, months or years** Age (number)____________________*(number box)* Days, weeks, months or years_______________*(drop down box, days [l-31]/weeks [l-52]/months [l-48]/years [1 W])*
6. **Please select any additional conditions that your child currently has. Please select all that apply**
  1. Continuous spikes and waves during slow-wave sleep syndrome (CSWS)
  2. Electrical Status Epilepticus during Slow Sleep (ESES/ESESS)
  3. Infantile spasms
  4. Movement disorder - ataxia, paroxysmal dyskinesia, dystonia, choreoathetosis
  5. Hypotonia
  6. Autism
  7. Intellectual disability
  8. Difficulty swallowing
  9. Uses a gastric tube for feeding and medicines
  10. Requires oxygen/ventilation support
  11. Visual impairment such as cortical blindness
  12. Variants in additional genes (please specify)____________(free *text box)*
  13. Other (please specify)_______________*(free text box)*
  14. No additional conditions
7. **What is the name and location of the hospital(s)/institution(s) where your child sees their neurologist/epileptologist? To protect their privacy, please do not provide the name of the physician** Name of hospital______________*(free text box)* Location of hospital__________________*(free text box)* Name of additional hospital_____________________*(free text box* Location of additional hospital________________*(free text box)*

#### SEIZURE HISTORY

8. **Has your child ever experienced seizures?** *(select one)*
  1. Yes *(answer question 9)*
  2. No *(skip to question 31)*
9. **Approximately how old was your child when they had their FIRST seizure? Please provide your best estimate in days, weeks, months, or years** Age (number)___________________*(number box)* Days, weeks, months or years___________________*(drop down box: days [0-31]/weeks [l-52]/months [l-48]/years [160])*
10. Approximately how often were your child’s seizures when they FIRST started, before a diagnosis of epilepsy was made? Please provide your best estimate *(select one)*
  1. More than 10 times per day
  2. Between 2-10 times per day
  3. Once per day
  4. Once per week
  5. Once per month
  6. Less than once per month
  7. Other (please specify)______________*(free text box)*
11. **What were your child’s seizure types when they FIRST started? Please select all that apply**
  1. Generalized tonic-clonic seizures
  2. Atonic seizures (drop seizures)
  3. Absence seizures (staring spells)
  4. Myoclonic seizures (shaking only without loss of consciousness)
  5. Partial/focal seizures
  6. Infantile/epileptic spasms
  7. Other (please specify)________________*(free text box)*
  8. Don’t know/Unsure
12. **Approximately how often does your child CURRENTLY have seizures? Please provide your best estimate** *(select one)*
  1. More than 10 times per day
  2. Between 2-10 times per day
  3. Once per day
  4. Once per week
  5. Once per month
  6. Less than once per month
  7. They are seizure-free *(skip to question 14)*
  8. Other (please specify)________________*(free text box)*
13. **What seizure type does your child CURRENTLY experience? Please select all that apply**
  1. Generalized tonic-clonic seizures
  2. Atonic seizures (drop seizures)
  3. Absence seizures (staring spells)
  4. Myoclonic seizures (shaking only without loss of consciousness)
  5. Partial/focal seizures
  6. Infantile/epileptic spasms
  7. Other (please specify)________________*(free text box)*
  8. Don’t know/Unsure
14. **Does your child experience seizures in ’dusters’? A seizure duster here is defined as multiple or repetitive seizures that stop and start, but occur in groups or clusters, quickly after one another** *(select one)*
  1. Yes, they usually occur in clusters (answer question 15A)
  2. Yes, they sometimes occur in clusters (answer question ISA)
  3. Yes, they used to experience seizure dusters, but not now (skip to question 15B)
  4. No, they haven’t experienced seizure dusters (skip to question 16)
15. **(A) Approximately how often does your child CURRENTLY experience seizure clusters? Please provide your best estimate** *(select one)*
  1. Once per day
  2. Once per week
  3. Once per month
  4. Once every 3 months
  5. Once per year
  6. Other (plea se specify)__________________*(free text box)*
15. **(B) Approximately how often did your child experience seizure clusters? Please provide your best estimate** *(select one)*
  1. Once per day
  2. Once per week
  3. Once per month
  4. Once every 3 months
  5. Once per year
  6. Other (please specify)__________________*(free text box)*
16. **When your child’s seizures were at their absolute WORST, approximately how often was your child experiencing seizures? Please provide your best estimate** *(select one)*
  1. More than 10 times per day
  2. Between 2-10 times per day
  3. Once per day
  4. Once per week
  5. Once per month
  6. Less than once per month
17. **Approximately how old was your child when their seizures were at their absolute WORST?** Age (number)______________*(number box)* Days, weeks, months or years________________*(drop down box: days [0-31 J/weeks [l-52]/months [l-48]/years [1 60])*
18. **For approximately how long were your child’s seizures at their absolute WORST before things started to improve? Please provide your best estimate. If you feel that their seizures are still at their absolute WORST, please provide an estimate to present date andcheckthe box below** Length of Time (number)_______________*(number box)* Days, weeks, months or years____________*(drop down box: days [0-31]/weeks [l-52]/months [l-48]/years [1 10])* My child’s seizures are still at their absolute worst______________________ *(check boxf(optional)*
19. **Please describe in your own words how your child’s condition changed overall when their seizures were at their absolute WORST** *(free text box)(optional)*
20. **Approximately how many seizures has your child had during the following time periods? This maybe challenging, but please provide your best estimate** Previous 1 month______________*’number box)* Previous 3 months______________*(number box)* Previous 6 months________________*(number box)*
21. **In the past 6 months, what is the longest period your child has been seizure free? Please provide your best estimate in days, weeks or months** Number_______________*(number box)* Days, weeks or months______________*(drop down box: days [0-186]/weeks [0-26]/months [0-6])*
22. **Approximately how long ago was your child’s last seizure?** Number*(number box)* Days, weeks, months or years*(drop down box: days [0-365]/weeks [0-52]/months [0-48]/years [0-60])*
23. **Overall, how would you describe the progression of your child’s seizures since onset?** *(select one)*
  1. Improved over time
  2. Unchanged
  3. Worsened over time
  4. Variable - have been periods of worsening and/or improvement
24. **Have you ever been trained to count or identify your child’s seizures?** *(select one)*
  1. Yes (answer question 25)
  2. No (skip to question 26)
25. **How was this training provided?**_______________________*(free text box)*
26. **On a scale of 1-10, how confident are you that you are able to identify your child’s seizures?** *(Select one)*

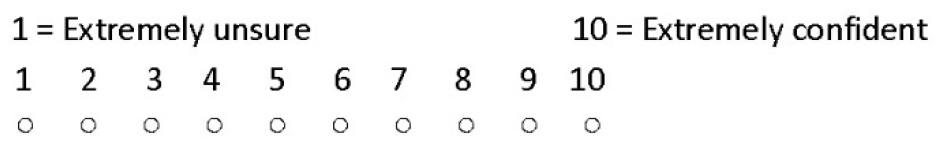
27. **Please feel free add any other information that you feel is important about the onset, frequency and severity of your child’s seizures***(free text box)(optional)*

#### EIZURE TREATMENT

26. **Please list the daily anti-epilepsy drugs (AEDs) or other treatments and supplements that your child is currently taking, or has taken previously, to control seizures: Please select all that apply** *(table)*

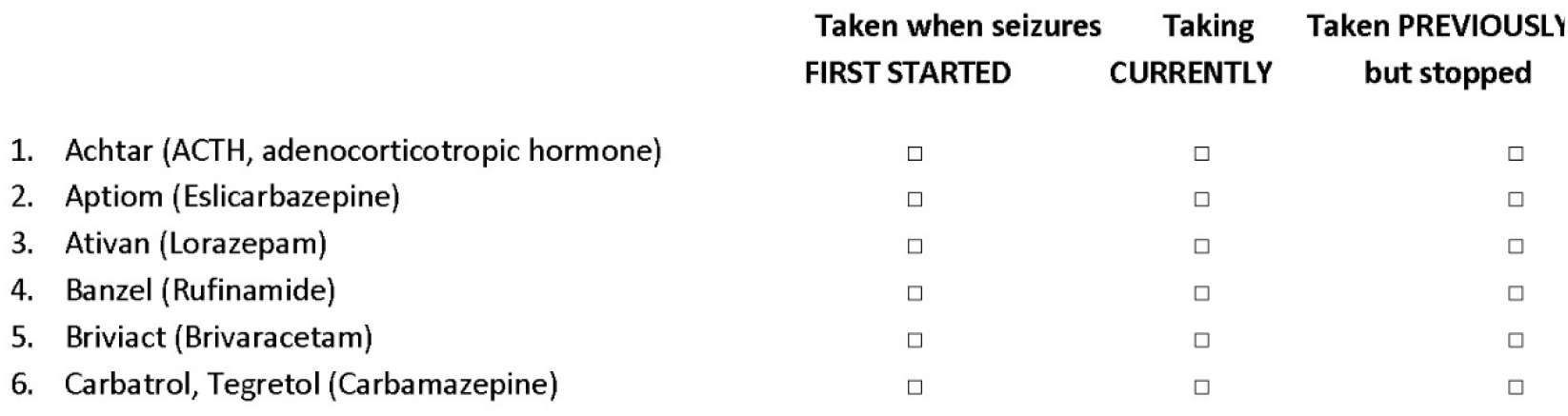
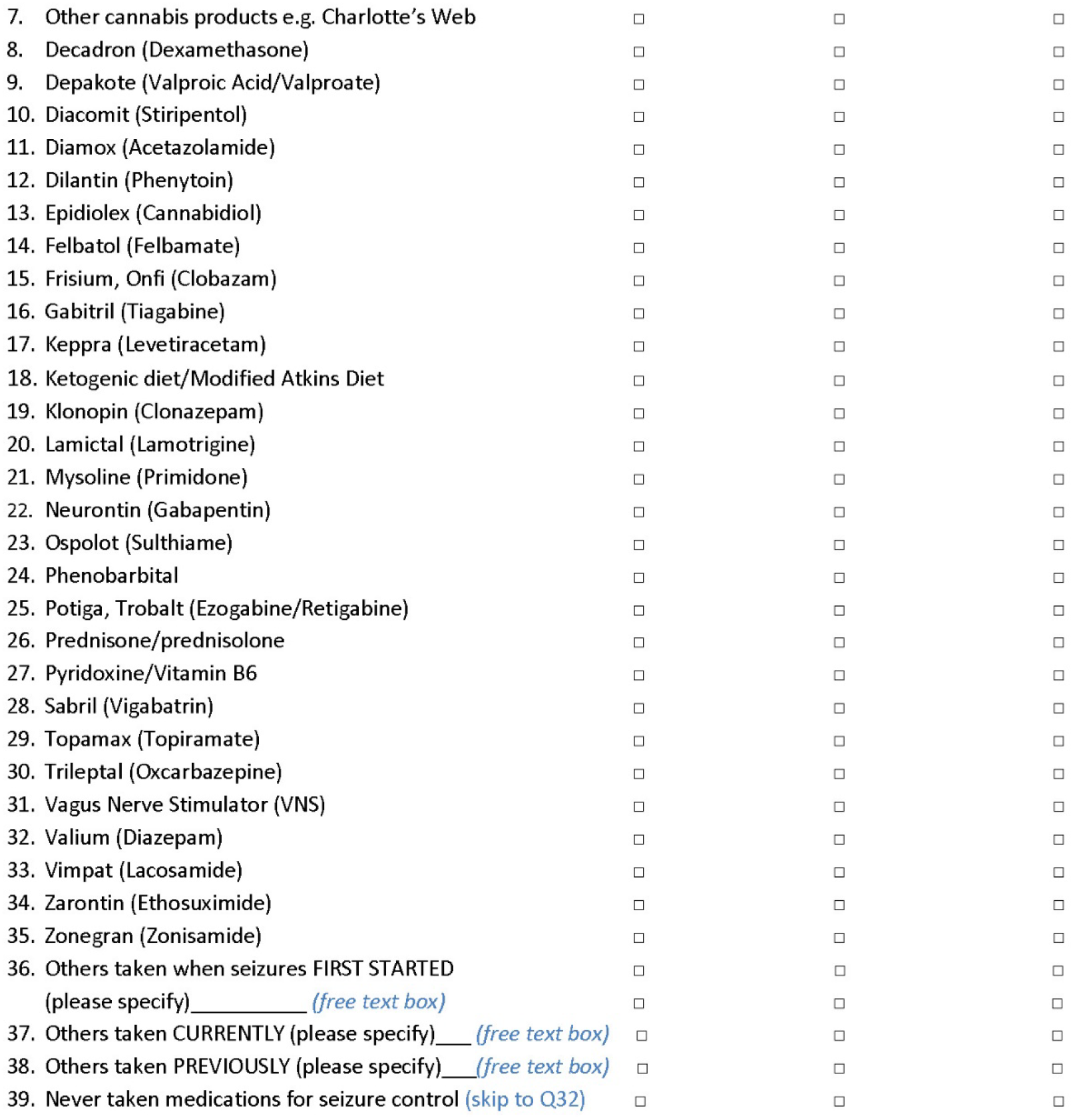
29. **Of the medications or treatments that your child has used, which do you believe have been the BEST at controlling your child’s seizures and why?***(free text box)*
30. **Of the medications or treatments that your child has used, which do you consider to be the WORST for controlling your child’s seizures and why?***(free text box)*
31. **Of the medications or treatments that your child has used, which do you consider to be the BEST for your child’s overall quality of life and why?***(free text box)*

#### FUTURE CLINICAL TRIALS

32. **What do you think are the most important things for new treatments or therapies to improve for your child and why?***(free text box)*
33. **Would you be willing for your child to participate in a clinical trial for a new drug to treat your child’s condition?** Please note this is to explore your potential level of interest, and does not mean that you are signing up for a trial *(select one)*
  1. Yes (skip to question 35)
  2. No (answer question 34)
34. **If no, please state why**_______________*(free text box)*
35. **Would you be willing to participate in further Xenon-sponsored surveys relating to other aspects of your child’s condition such as neurodevelopment and quality of life or the potential design of a clinical trial?** *(select one)*
  1. Yes, please enter your email address:*(email or free text box)*
  2. No
36. **How did you find out about this survey? Please select all that apply**
  1. The Cute Syndrome Foundation website
  2. Email from The Cute Syndrome Foundation
  3. Facebook
  4. Webinar
  5. From a friend or family member
  6. Other (please specify)*(free text box)*

Thank you for completing the Survey.

**Supplementary Table 1.** Free-text Response Reasons for Stopped or Worst Treatments^a^.

|  | Levetiracetam | Topiramate | Phenytoin | Lacosamide | Clobazam | Valproate | Cannabidiol | Oxcarbazepine | Vigabatrin | Carbamazepine | Phenobarbital |
| --- | --- | --- | --- | --- | --- | --- | --- | --- | --- | --- | --- |
| Total responses | 56 | 12 | 7 | 6 | 5 | 5 | 4 | 4 | 4 | 3 | 3 |
| Increase in seizure frequency | 13 | 2 | 0 | 1 | 0 | 1 | 0 | 1 | 1 | 0 | 0 |
| Increase in seizure severity | 17 | 2 | 2 | 1 | 1 | 1 | 1 | 1 | 2 | 2 | 0 |
| Change in seizure type | 4 | 1 | 0 | 0 | 0 | 1 | 0 | 0 | 0 | 0 | 0 |
| Increase in duration of seizure | 0 | 0 | 0 | 1 | 0 | 0 | 0 | 0 | 0 | 0 | 0 |
| No improvement | 14 | 3 | 2 | 1 | 2 | 0 | 1 | 1 | 1 | 0 | 3 |
| Any side effect | 7 | 4 | 3 | 2 | 2 | 2 | 2 | 1 | 0 | 1 | 0 |
| Common side effects<sup>b</sup> |  |  |  |  |  |  |  |  |  |  |  |
| GI problems<sup>c</sup> | 0 | 1 | 0 | 0 | 0 | 0 | 2 | 0 | 0 | 1 | 0 |
| Behavioral problems/negative behavioral side effects | 3 | 0 | 1 | 0 | 0 | 0 | 0 | 0 | 0 | 0 | 0 |
| Developmental delays | 0 | 2 | 0 | 0 | 0 | 0 | 0 | 0 | 0 | 0 | 0 |

<sup>a</sup> Includes ASMs with ≥3 responses. Caregivers could report >1 reason for stopping treatment if needed.
<sup>b</sup> Reported in ≥2 patients.
<sup>c</sup> Includes the responses “nausea/vomiting”, “losing all ability to hold food down”, “major GI side effects”, “extremely suppressed appetite”, and “constipation”. ASM, anti-seizure medicine; GI, gastrointestinal.

